# WOMEN’S BIRTH PREPAREDNESS AND COMPLICATION READINESS IN NIGERIA: A SYSTEMATIC REVIEW AND META-ANALYSIS

**DOI:** 10.1101/2021.11.03.21265854

**Authors:** Margaret O. Akinwaare, Abimbola Oluwatosin, Olakekan Uthman, Elizabeth Ike

## Abstract

**Introduction:** Globally, efforts are being made to reduce the menace of maternal death in order to achieve the sustainable development goal. Maternal death has been associated with inadequate birth preparedness and complication readiness especially in the low and middle-income countries. Therefore, this review assessed birth preparedness and complication readiness in Nigeria.

**Methods:** A systematic review and meta-analysis of published research articles on birth preparedness and complication readiness in Nigeria was done using PubMed, EMBASE and MEDLINE databases. All published articles from inception to November, 2018 were included in the review. A total of 8913 published articles were identified from electronic search, a total of 4440 studies were included in this review, while only 12 articles met the inclusion criteria and were included in the meta-analysis.

**Results:** The pooled prevalence of ‘Good BPCR’ for all studies yielded an estimate of 58.7% (95% CI 43.9 to 72.7%). The *I*^*2*^ statistic was 98%, indicating statistically significant heterogeneity among the studies. The percentage of women with good birth preparedness and complication readiness increases with increasing year of publication, such that women tended to be more aware good birth preparedness and complication readiness in recent years. More than half of the women had knowledge of obstetric danger signs (52.0%, 95% CI 39.5 to 64.4%, 10 studies), arranged for transportation (59.5%, 95% CI 36.2 to 80.7, 11 studies) or saved money (63.4%, 95% CI 44.7 to 80.2%, 11 studies) as part of the BPCR.

**Conclusion:** Women in Nigeria are better prepared for birth preparedness and complication readiness in recent years. Therefore, interventions to promote more adequate birth preparedness and complication readiness among women is recommended.

**PROSPERO registration number:** The study protocol was registered with PROSPERO number, CRD42019123220

**Key questions:** *What is already known?:* Globally, birth preparedness and complication readiness is a concept which has been proven to be effective at increasing skilled birth attendance. It is a strategy for reducing maternal death especially in middle and lo-income countries.

*What are the new findings?:*

- More pregnant women and recently delivered mothers in the Southern part of Nigeria have a better birth preparedness and complication readiness compared to their counterpart in the Northern part of the country.
- Number of pregnant women and recently delivered mothers with good birth preparedness and complication readiness increases with increasing year of publication.

*What do the new findings imply?:*

- This review provides data about women’s birth preparedness and complication readiness across different regions of Nigeria. Having a poorer BPCR in the Northern region of the country could be responsible for higher maternal death in the region.
- This could be used to plan interventions to improve birth preparedness and complication readiness in different regions of the country.

*Data availability statement:* No data are available.

## Background

The World Health Organization (WHO) has a vision of universal coverage of health care which corroborates the third Sustainable Development Goal (SDG) - to ensure healthy lives and promote wellbeing for all at all ages. However, due to disparity in the health systems, the lifetime risk of maternal mortality in the sub-Saharan region is estimated to be 1 in 38^1^, while is estimated to be 1 in 3,700 in the developed world. Pregnant women in the developing countries like Nigeria have a high risk of mortality^2^. In Nigeria, the maternal mortality ratio (MMR) was at 814 per 100,000 live births as at 2015^3^ and the annual maternal death was 40,000 which contributes 14% to the global aggregate^2^. In spite of this unacceptably high MMR, there are proven interventions that are known to prevent maternal deaths. These include birth preparedness and complication readiness resulting in skilled attendance for pregnancy and childbirth as well as access to emergency obstetric care.

Inadequate BPCR is one of several factors contributing to maternal deaths while adequate BPCR could determine the survival of a pregnant woman and her unborn child^4^. Birth and emergency preparedness is an integral component of antenatal care which involves planning with the key stakeholders; the healthcare provider, pregnant women, relatives and the community^5^. BPCR is a key component of globally accepted safe motherhood programs which helps women reach out for professional delivery care when labour begins. This in turn reduces delays that occur when mothers in labour experience obstetric complications^6^. The BPCR plan encourages women, households, and communities to make arrangements such as identifying or establishing available transport, setting aside money to pay for service fees and transport, and identifying a blood donor in order to facilitate swift decision-making and reduce delays in reaching care once a problem arises.

One of the major strategies to reduce maternal mortality is making plans for both normal birth and any unpredicted complication that may arise from pregnancy to childbirth (BPCR). According to Ayuba, et al^7^, BPCR is defined as “a set of knowledge, behaviours and actions undertaken by women, families, communities, health care providers and facilities to enhance the survival of women and new-borns during pregnancy, childbirth and the postpartum period”. This is corroborated by Ekabua et al^8^, who documented BPCR as a process of planning for normal birth and anticipating the actions needed in case of an emergency. It is the advanced planning and preparation for delivery in order to improve maternal and new-born health outcomes.

This systematic review and meta-analysis provide an assessment of BPCR strategies as it relates to reduction of maternal and child mortality and the most effective components of BPCR.

## Objective of the review

The primary aim of this review is to assess birth preparedness and complication readiness among pregnant women and recently delivered women in Nigeria.

## Methods

### Study design and search strategy

A systematic review and meta-analysis of published research articles on birth preparedness and complication readiness in Nigeria was done. PubMed, EMBASE and MEDLINE databases were searched using the medical subject heading (MeSH) terms and the following search terms: Birth, “Birth preparedness”, “Complication readiness”, “Birth preparedness” AND “complication readiness”, “Birth preparedness” OR “complication readiness”, Birth AND “emergency preparedness”, Nigeria. All published articles from inception to November, 2018 were included in the review.

### Study selection

The inclusion criteria:

1. **Type of study**: the review included observational and experimental studies on birth preparedness and complication readiness
2. **Settings:** the review included studies conducted at urban and rural areas irrespective of whether the study is hospital-based or community-based.
3. **Participants**: the review included studies conducted on birth preparedness and complication readiness among pregnant women and recently delivered women in Nigeria.
4. **Outcome**: the review included studies which reported knowledge and practice of birth preparedness and complication readiness as primary and/or secondary outcome.
5. **Time frame**: studies from inception to November, 2018 were included in the review.
6. **Language:** only studies published in English language were included in the review
7. **Publication type**: journal articles

Exclusion criteria include:

1. Studies that failed to list elements of birth preparedness and complication readiness measured to conclude on its practice was excluded from the review.
2. Studies which did not specify the study participants will be excluded from the review.

### Selection process

The full text of all the studies which met the stated inclusion criteria were retrieved and reviewed by two reviewers independently. The two reviewers independently evaluated the eligibility of the studies obtained from the literature searches. All articles yielded by the database search was initially screened by their titles and abstracts to obtain studies that met inclusion criteria. The generated reports from the two reviewers were retrieved. In cases of discrepancies, agreement was reached by consensus.

### Data extraction and data collection

Data extraction tool was prepared for this study. The tool includes information on study design, study area, sample size, participants, publication year, knowledge of birth preparedness and complication readiness, practice of birth preparedness and complication readiness, knowledge of obstetric danger signs, percentage of women who saved money, percentage of women who arranged for transportation, percentage of women who identified/arranged for compatible blood donor, percentage of women who identified skilled birth attendants, percentage of women who identified place of birth, percentage of women who arranged for companion to health facility during labor or any emergency and percentage of women who arranged for whom to care for the family while away for delivery or any emergency.

### Main outcome(s)

#### Primary outcome

Birth preparedness in accordance with Johns Hopkins Program for International Education in Gynaecology and Obstetrics (JHPIEGO, 2004):

- Choosing of birth location
- Arrangement for transportation to skilled care site
- Obtaining basic birth supplies
- Saving money for skilled care at childbirth

Complication readiness: in accordance with Johns Hopkins Program for International Education in Gynaecology and Obstetrics (JHPIEGO, 2004):

- Good knowledge of obstetric danger signs
- Saving money for skilled emergency care
- Identifying a compatible blood donor
- Arrangement for transportation in case of emergency
- Arrangement for household support in case of emergency.

We will exclude studies that failed to list elements of BPCR measured to conclude on BPCR practice.

#### Secondary outcomes

- Knowledge of obstetric danger signs

#### Risk of bias (quality) assessment

Risk of Bias Assessment tool for Non-randomized Studies (RoBANS) and “A Cochrane Risk Of Bias Assessment Tool for Non-Randomized Studies” (ACROBAT-NRSI) were adapted to appraise the risk of bias for the included studies. (see Box 1). The risk of bias was assessed by scoring (low risk = 1, unclear = 0, high risk = -1) each bias type for each publication and the total score was used as the summary assessment of risk of bias.

##### Box 1

**Risk of bias assessment**

|  |
| --- |
| **Box 1: Risk of bias assessment** |

| **Bias type** | **Low-risk of bias** | **High-risk of bias** | **Unclear risk of bias** |
| --- | --- | --- | --- |
| Selection (sample population) | participants selected randomly | Sample selection ambiguous and sample unlikely to be representative | Insufficient information |
| Selection (participation rate) | High participation rate (>70-85%) | Low participation rate (<70%) | Insufficient information |
| Performance bias (outcome assessment) | Objective measures of BPCR | Self-reported measure of BPCR | Insufficient information |
| Performance bias (analytical methods to control for bias) | Analysis appropriate for type of sample (unadjusted, univariable analyses etc.) | Analysis does not account for common adjustment (adjusted, multivariable analyses) | Insufficient information |
| Other form of bias | There is no evidence of bias from other sources. | There is potential bias present from other sources | Insufficient information |

For each included study, we estimated the precision (C) or margin of error, considering the sample size (SS) and the observed BPCR practice proportion among pregnant women and recently delivered mothers from the formula:

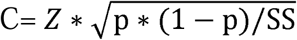

where Z was the z-value fixed at 1.96 across studies (corresponding to 95% confidence interval). The desirable margin of error is 5% (0.05) or lower.

#### Synthesis of results

We pooled the prevalence estimates using the DerSimonian–Laird random effects model^9.^ We assessed the between study variations in prevalence estimates using the Higgins *I*^*2*^ statistic, and, as recommended, a value of greater than 50% was considered for moderate heterogeneity^10,11^. We conducted series of subgroup analyses to explore the effect of study-level covariates on the prevalence estimates^12^. The possibility of reporting bias was assessed using an Begger’s test and funnel plot^13^. Leave-one-study-out sensitivity analysis was also performed to assess the stability of the meta-analysis result^14^. Stata 16 was used for all analyses (StataCorp, College Station, TX).

#### Ethical consideration

This study followed the registered protocol.

#### Patient and public involvement

Neither patients nor the public were involved in the study design and article selection.

## Results

### Studies included in the meta-analysis

A total of 8913 published articles were identified from electronic search; 8830 from Google scholar, 71 from PubMed and 12 from MEDLINE. However, 23 articles were excluded for not reporting outcome variables, 63 were excluded for duplication, 7921 were excluded through review of titles, while 894 were excluded through reviewing of abstract. Hence, 12 articles met the inclusion criteria and were included in the meta-analysis (Figure 1). However, the total sample size of included studies in this review was 4440; 1390 from the south-west, 1443 from the south-east, 1207 from the south-south and 400 from the north-west (Table 1).

**Figure 1:**
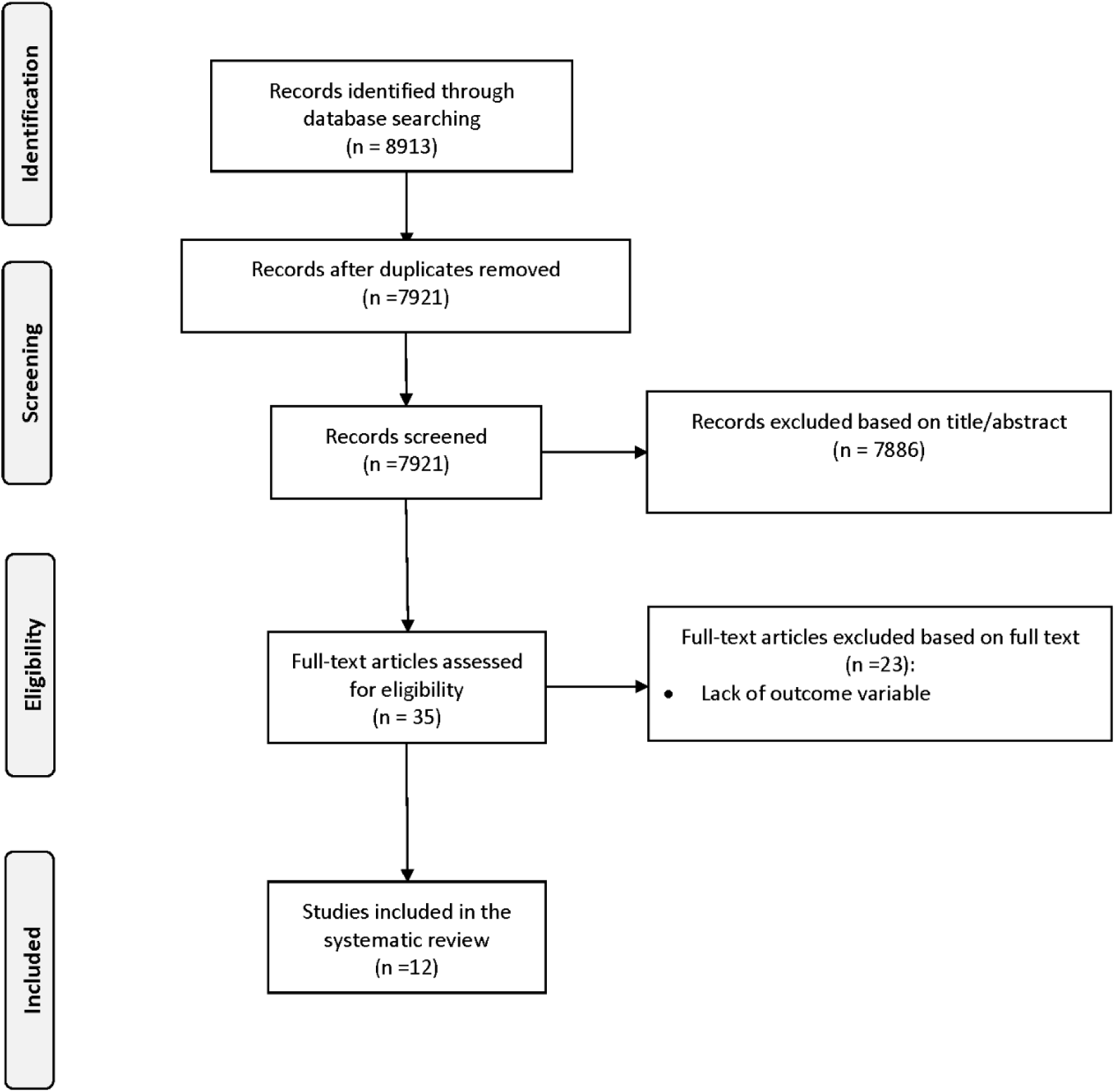
Flow diagram of studies included in analysis.

**Table 1:** Characteristics of birth preparedness and complication readiness studies included in meta-analysis.

| S/N | Authors | Study design | Study area | Study year | Sample size | Population | % of good BPCR |
| --- | --- | --- | --- | --- | --- | --- | --- |
| 1 | Onayade et al<sup>4</sup> | Cross-sectional | Ile-Ife, Osun state | 2010 | 400 | Pregnant women | 61% |
| 2 | Illiyazu et al<sup>25</sup> | Cross-sectional | Ungogo, Kano state | 2010 | 400 | Married men and their wives | 32% |
| 3 | Ekabua et al<sup>8</sup> | Cross-sectional | Calabar,<br>Cross<br>River state | 2011 | 800 | Recently<br>delivered<br>mothers | 77% |
| 4 | Tobin et al<sup>26</sup> | Cross-sectional | Oredo,<br>Benin city,<br>Edo state | 2014 | 230 | Pregnant<br>women | 87% |
| 5 | Emma-Ukaegbu et<br>al<sup>27</sup> | Cross-sectional | Umuahia,<br>Abia state | 2014 | 474 | Pregnant<br>women | 33% |
| 6 | Idowu et al<sup>28</sup> | Cross-sectional | Ogbomoso,<br>Oyo state | 2015 | 400 | Pregnant<br>women | 40% |
| 7 | Oni et al<sup>29</sup> | Cross-sectional | Lagos,<br>Lagos state | 2016 | 410 | Pregnant<br>women | 22% |
| 8 | Ibadin et al<sup>30</sup> | Cross-sectional | Anegbette,<br>Etsako,<br>Edo state. | 2016 | 277 | Pregnant<br>women | 48% |
| 9 | Obi et al<sup>31</sup> | Cross-sectional | Benin city,<br>Edo state | 2016 | 252 | Pregnant<br>women | 78% |
| 10 | Sabageh et al<sup>32</sup> | Cross-sectional | Osogbo,<br>Osun state | 2017 | 180 | Pregnant<br>women | 82% |
| 11 | Nkwocha et<br>al<sup>33</sup> | Cross-sectional | Port-Hartcourt,<br>Rivers<br>state. | 2017 | 407 | Pregnant<br>women |  |
| 12 | Ohamaeme et<br>al<sup>34</sup> | Cross-sectional | Imo state | 2017 | 210 | Pregnant<br>women | 77% |

### Overall prevalence of Birth preparedness and complication readiness in Nigeria

The percentage of women with good birth preparedness and complication readiness and 95% CIs from individual studies with a pooled estimate are shown in Fig. 2. The percentage of women with good birth preparedness and complication readiness ranged from as low as 22.0% to as much as 87.0%. The pooled prevalence of ‘Good BPCR’ for all studies yielded an estimate of 58.7% (95% CI 43.9 to 72.7%). The *I*^*2*^ statistic was 98%, indicating statistically significant heterogeneity among the studies. The contour-enhanced funnel plot of examination of publication bias is shown in Fig. S1 in the online data supplement. We found no evidence of publication bias as indicated by the relatively symmetrical funnel plot of studies’ precision against prevalence estimates (in logarithmic scale). This was confirmed when formally tested using the Begg’s method (p value for small study bias□=□0.2129). The results of leave-one-study-out sensitivity analyses showed that no study had undue influence on the pooled prevalence estimates.

**Figure 2:**
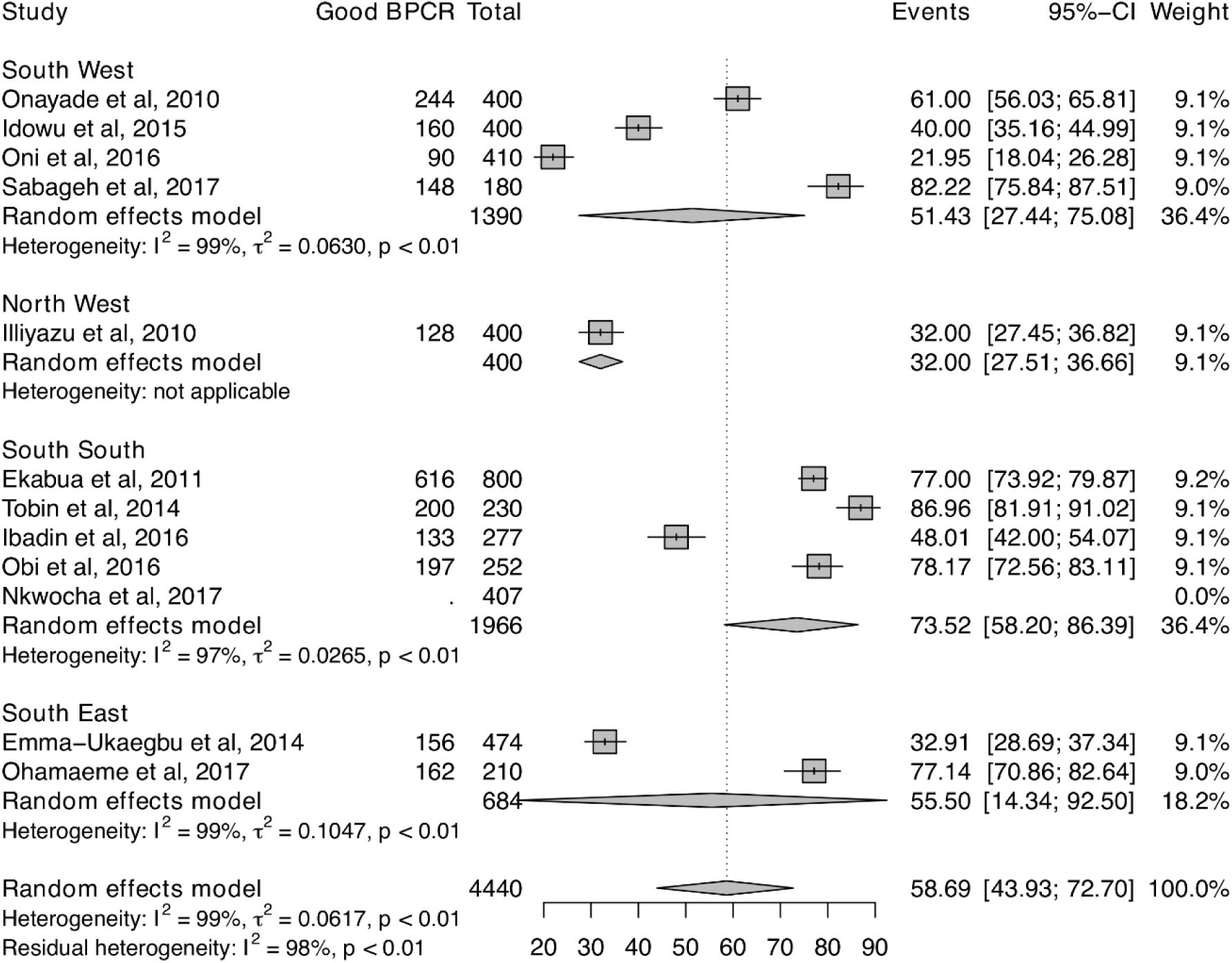
Forest plot showing percentage of women with good birth preparedness and complication readiness studie.

As shown in Fig. 2 The percentage of women with good birth preparedness and complication readiness tended to be higher among women from the Southern part of Nigeria than those from the Northern part of the country. In addition, we found evidence that the percentage of women with good birth preparedness and complication readiness increases with increasing year of publication, such that women tended to be more aware good birth preparedness and complication readiness in recent years (Figure 3). Furthermore, we found evidence of small study bias, i.e. the percentage of women with good birth preparedness and complication readiness tended among studies with small sample sizes (Figure 4).

**Figure 3:**
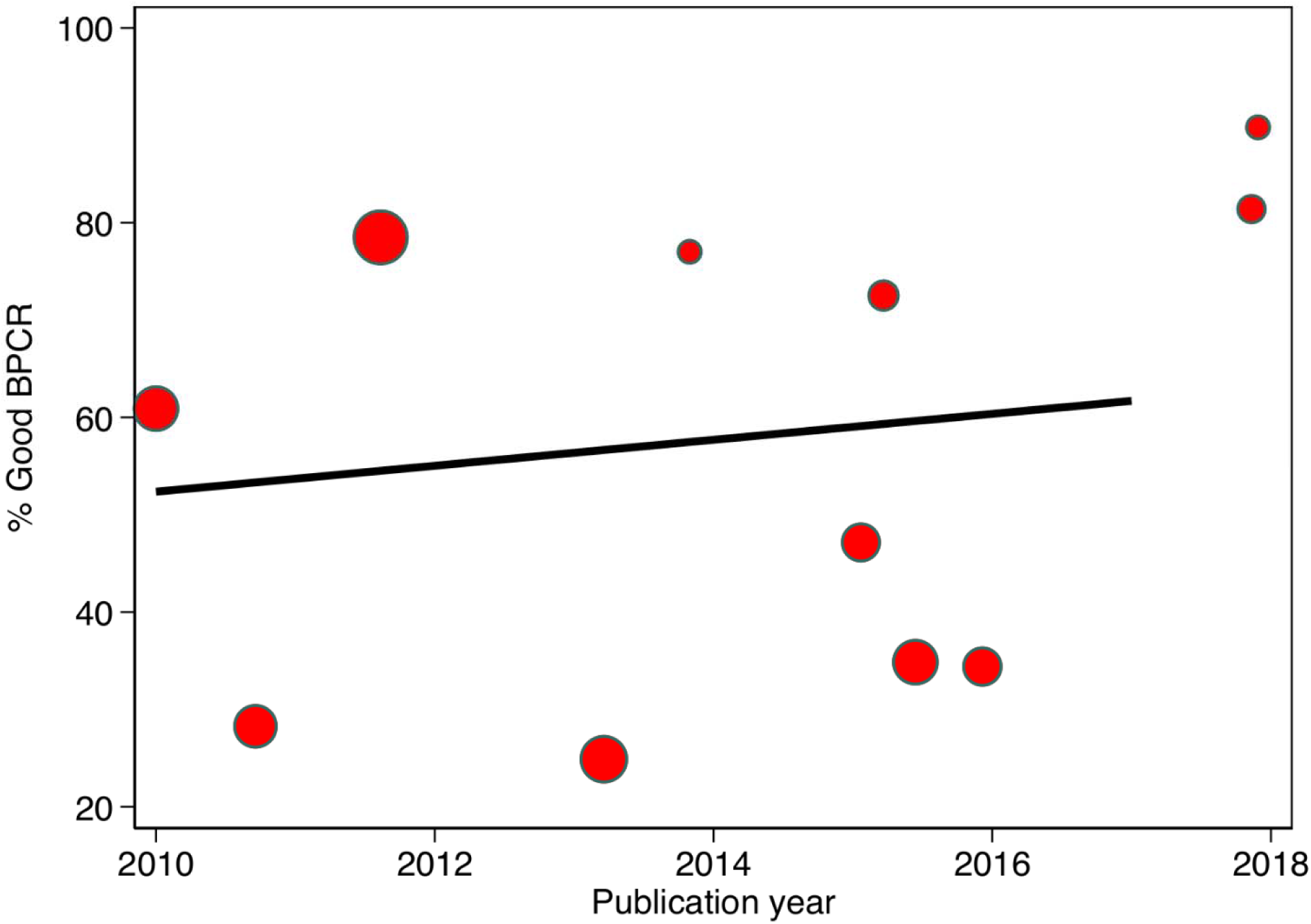
Secular trends in percentage of women with good birth preparedness and complication readiness studies.

**Figure 4:**
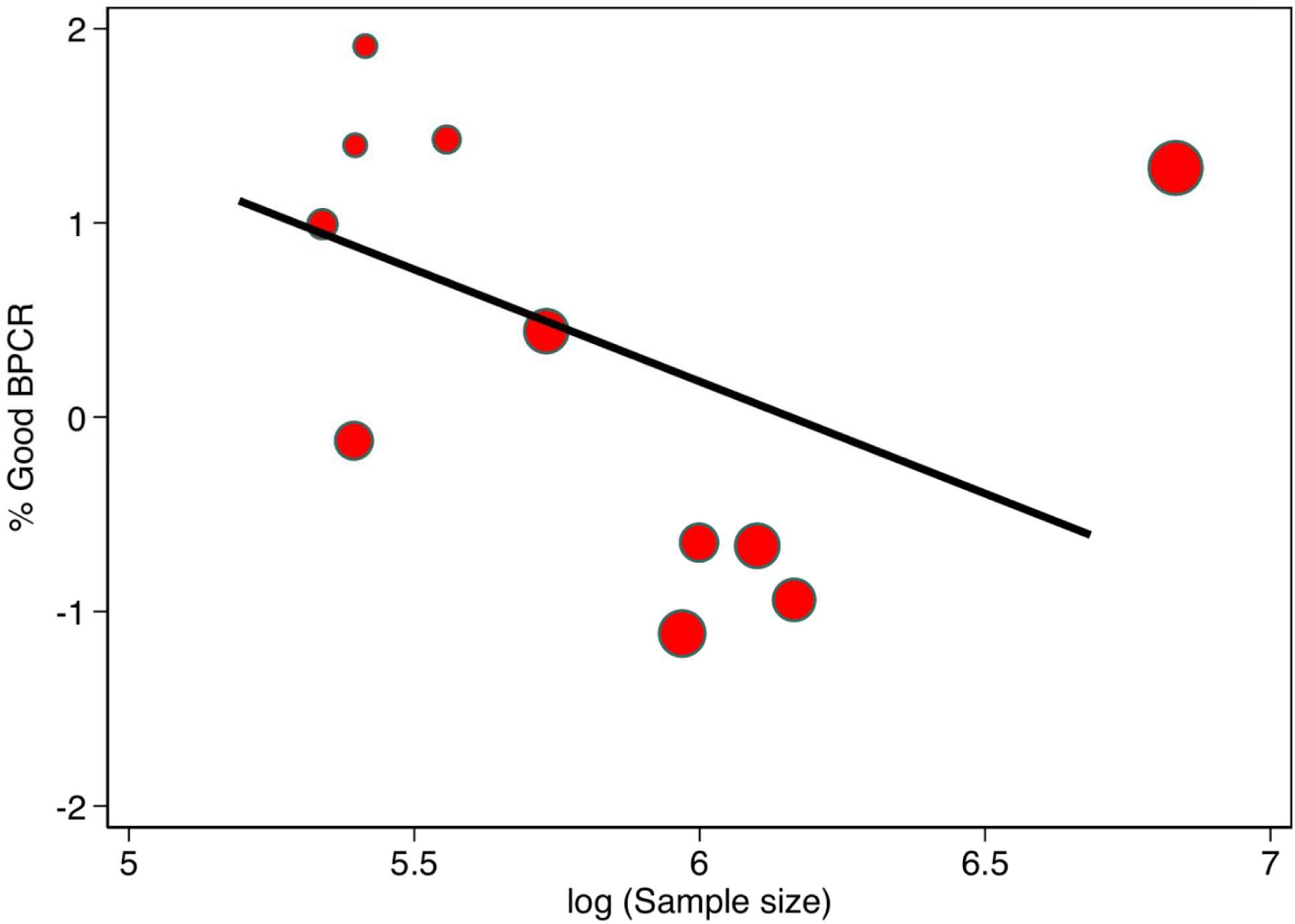
Small study effects.

### Other components of birth preparedness and complication readiness in Nigeria

The results of other components of birth preparedness and complication readiness in Nigeria are shown in Fig. 5. On average, preponderance of the women obtained supplies as part of the BPCR (77.1%, 95% CI 56.5 to 92.5%, 8 studies). About 7 in 10 women identified skilled birth attendance (69.7%, 95% CI 45.7 to 89.1%, 8 studies) or identified place of birth as part of the BPCR (69.9%, 95% CI 40.2 to 92.5%, 8 studies) as part of the BPCR. More than half of the women had knowledge of obstetric danger signs (52.0%, 95% CI 39.5 to 64.4%, 10 studies), arranged for transportation (59.5%, 95% CI 36.2 to 80.7, 11 studies) or saved money (63.4%, 95% CI 44.7 to 80.2%, 11 studies) as part of the BPCR. Less than 50% of the women arranged for companion to healthcare facility as part of BPCR (47.1%, 95% CI 24.5 to 70.3%, 5 studies). On average only about 16.7% (95% CI 6.35 to 30.72%, 11 studies) of the women arranged for blood donor as part of the BPCR.

**Figure 5:**
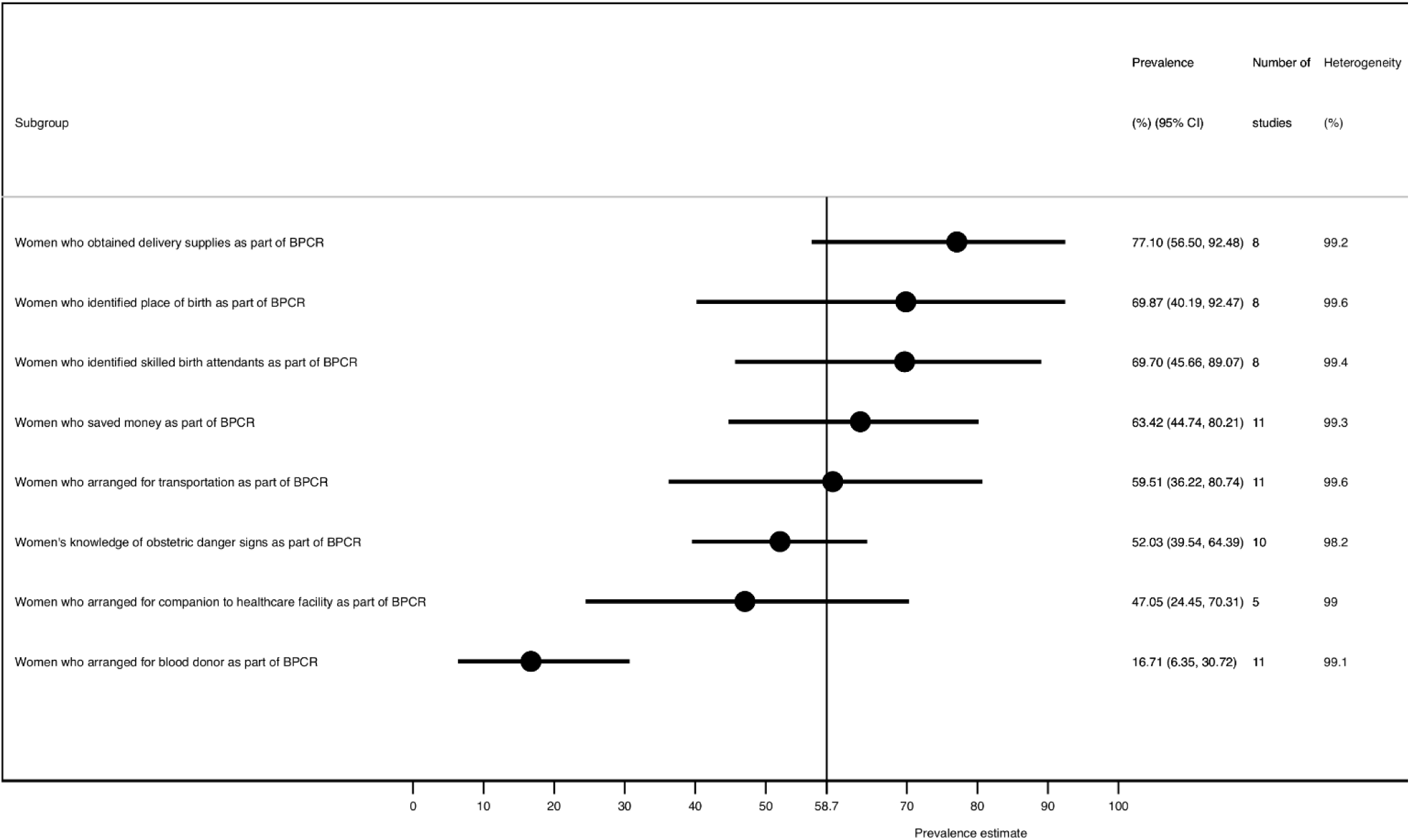
Other components of birth preparedness and complication readiness in Nigeria.

## Discussion

The aim of this systemic review and meta-analysis was to assess birth preparedness and complication readiness among pregnant women and recently delivered women in Nigeria. The total sample size of included studies in this review was 4440 while twelve studies met the inclusion criteria and were included in the meta-analysis.

The components of birth preparedness and complication readiness identified in this review is similar to that of the review conducted in Ethiopia^15^. Findings of the meta-analysis revealed that more than half of the women had good birth preparedness and complication readiness. This is similar to a study conducted in West Bengal, India^16^ and Central Tanzania^17^. However, the outcome in this review is better compared with the meta-analysis carried out in Ethiopia which reported that only one-thirds of pregnant women in Ethiopia were prepared for birth and its complication^15^. Also, it is higher than the studies carried out in Uganda, Bangladesh^18^ and Gambia^19^. In contrast, it is lower than the studies conducted in Accra^20^.

The discrepancies may probably be due to the local context like priority of government on implementation of safe motherhood programmes including BPCR. In the context of this review the level of good birth preparedness and complication readiness is commendable, however, a better result was expected considering the Nigerian government and non-governmental organizations policies and initiatives on maternal mortality reduction in Nigeria using birth preparedness and complication readiness as a strategy.

Also, it was observed in this review that the percentage of women with good birth preparedness and complication readiness tended higher among women from the Southern part of Nigeria than those from the Northern part of the country. This could be a probable explanation of the high maternal mortality in the northern region^21,22^. The reason could be socio-cultural issues like religion, educational status and economic status of the women. In the northern part of Nigeria, they practice Islamic culture referred to ‘pudah’ which restrict women from being autonomous. The women are required to ask for their husbands’ permission when they need medical attention hence, they cannot plan or take decision concerning their health without the husbands’ approval. In addition, the Sharia law practiced in various states in the north deprive women of their rights to freedom of movement and association including access to education. In the light of this, most women in Northern Nigeria will neither utilize antenatal care in a healthcare facility nor embrace birth preparedness and complication readiness^23,24^.

## Data Availability

No data are available.

## Data availability statement

No data are available.

## Ethics statements

Patient consent for publication Not applicable.

## Registration and protocol

The study protocol was registered with PROSPERO number, CRD42019123220. No major amendment was made to information provided at registration or in the protocol. The protocol can be accessed from: https://www.crd.york.ac.uk/prospero/display_record.php?ID=CRD42019123220

